# A NURSING PHILOSOPHY OF FAMILY EMPOWERMENT IN CARING UNDER-FIVE CHILDREN ILLNESS

**DOI:** 10.1101/2022.10.06.22280765

**Authors:** Praba Diyan Rachmawati, Yuni Sufyanti Arief, Moses Glorino Rumambo Pandin

## Abstract

**Introduction:** Sick children will be at risk of experiencing growth and development disorders and experiencing severe conditions to decreased quality of life. Quality of care through proper management of sick children under five is a priority. However, in the field of pediatric nursing, there are still obstacles to the application of family empowerment in caring for sick children, so it is important to examine a literature review with a philosophical approach the application of family empowerment in caring for sick children, as an effort to optimize the care of sick children under-five.

**Method:** This study was based on the results of the Literature Review. Articles were obtained from 3 databases, namely Scopus, Science Direct and PubMed. The keywords used in searching the literature in this study were ((parent) OR (mother) AND (parental AND empowerment) OR (engagement) AND (children) OR (sick AND children)). Articles searched from 2018-2022, which were open access and in English, from this literature review search, found 12 articles.

**Result:** Based on a philosophical approach of family empowerment in the care of sick children under-five, family empowerment interventions with the principle of involving families in care, and increasing family knowledge and skills in caring for sick children can be implemented as an effort to optimize care for sick toddlers.

**Conclusion:** The results of this literature review can be used as a basis for nursing interventions that require parental involvement in caring for sick children. Family empowerment programs that are planned and structured can be applied in the care of sick children at home or in the hospital.

## Introduction

Illness and mortality in children under five is a global problem that requires handling in various aspects holistically. Sick toddlers need serious attention, pain in toddlers if they don’t get prompt and proper treatment will fall into a severe condition that results in death (Rachmawati et al., 2022). the under-five mortality rate in Indonesia until 2017 was 32 per 1000 live births (USAID, 2020). Parents whose children are sick experience stressful conditions this is due to many factors, such as children’s health, financial problems, children’s behavior, and other environments, stress on parents often affects parenting behavior and child behavior (Damen et al., 2021). Families have a big role in child care, especially in the initial treatment of sick children or daily child care. The family’s ability to care for a sick child determines the severity or recovery of the child because the family is the unit closest to the child in daily life.

Children under five years of age are at the stage of the golden age period, so this toddler age is in a period of very rapid physical growth and development, the first 1000 days of a child’s life become a golden period which if passed with poor quality stimulation will have a negative impact on the life of the next child (Toto Sudargo, Tira Aristasari, 2018). Under-five children who are sick will experience a decrease in quality of life so they are at risk of experiencing obstacles to growth and development optimally, as well as families who care for children also experience a decrease in quality of life, due to anxiety, fatigue, helplessness, lack of knowledge and ability to care for children (Rachmawati et al., 2016).

The family is the smallest unit closest to the child, good family knowledge and skills in caring for sick toddlers determine the level of recovery. Research shows that parents need information from health workers to improve their skills in caring for sick children (May et al., 2018). Family Center Care improves child nursing services by involving parents in the stages of nursing care for sick children. Nurses have an important role in providing nursing care to sick toddlers, through family empowerment it is hoped that they can optimize the nursing interventions provided, so it is important to elaborate more deeply on family empowerment in a philosophical review of child nursing. This Literature review aims to describe family empowerment in caring for sick children based on a philosophical approach to ontology, epistemology and axiology.

## Method

This literature review is in the search for articles using the PICOS framework with the question “how is parental empowerment in treating sick children with a philosophical approach” taken from 3 data bases, namely Scopus, Science Direct and Pubmed which are limited from 2017-2022 and all articles are in English. The articles obtained were based on the keywords ((parent) OR (mother) AND (parental AND empowerment) OR (engagement) AND (children) OR (sick AND children)). All articles were analyzed by PRISMA Flowchart to get relevant articles.

**Table 1.** Characteristic of Reviewed Studied

| No | Author and Year | Title | Method | Results |
| --- | --- | --- | --- | --- |
| 1 | Arief, Y S, Nursalam, & Rachmawati, P. D (2020) | Family filial value in caring for children with leukemia.<br>Enfermeria Clinica | D: explanatory descriptive research<br>S: 15 families who have children with leukemia who are undergoing chemotherapy and mothers who take care of their children directly<br>V: Family values and family's ability to care for children with Leukemia<br>I: Filial score scale instrument and health status questionnaire<br>A: Descriptive analysis | Family empowerment in this study are divided into three factors. These factors are: responsibility in caring for children leukemia, respect for children, and care for caring for children suffering from leukemia |
| 2 | Damen, H., Scholte, R. H. J., Vermulst, A. A., van Steensel, P., & Veerman, J. W. (2021) | Parental empowerment as a buffer between parental stress and child behavioral problems after family treatment. | D: Pre-post test design<br>S: 275 Family<br>V: Children's behavioral problems, Parental Empowerment, Parental Stress, Professional support.<br>I: Parental Empowerment (EMPO), Parental Stress (Parenting Stress Questionnaire), SDQ | The level of parental empowerment after Intensive Family Treatment (IFT) has increased.<br>Parental stress has a bad influence on children's behavior |
| 3 | Krisnana, I., Sulistyarini, H., Rachmawati, P. D., Arief, Y. S., & Kurnia, I. D. (2019) | Reducing acute stress disorders in mothers of leukemic children by means of the family Centered Empowerment Module (FACE). | D: quasi-experimental pre-test/post-test control group<br>S: 60 Mothers with children with leukemia who are being treated at the hospital<br>V: Family Centered Empowerment Module (FACE), Stress on Mother<br>I: DASS and FACE Module | There was a decrease in the stress level of mothers with leukemia children in the intervention group after giving FACE |
| 4 | May, M., Brousseau, D. C., Nelson, D. A., Flynn, K. E., Wolf, M. S., Lepley, B., & Morrison, A. K (2018) | Why Parents Seek Care for Acute Illness in the Clinic or the ED: The Role of Health Literacy | D: Qualitative<br>S: Parents of children who are sick and visit the emergency department<br>V: Parent seek care<br>I : a semistructured interview guide | Factors that influence the behavior of parents in managing children with acute illness are literacy-related abilities, understanding of disease severity, ability to assess and treat illness at home and ability to seek health care systems. |
| 5 | Mohammadzadeh, E., Varzeshnejad, M., Masoumpour, | The impact of the family-centered | Design: Pre-test post test experimental study | FCEM is an effective intervention to reduce stress on parents and |
|  | A., & Ahmadimehr, F. (2022) | empowerment model on the children's quality of life with chemical burns and their parent's perceived stress | Variables: Quality of Life Children, Family-Centered Empowerment Model (FCEM)-based program<br>Instruments: PedsQL, TAPQOL<br>Analysis: The data were analyzed using SPSS statistical software V. 23. | improve the quality of life of children affected by burns |
| 6 | Piotrowska, P. J., Tully, L. A., Lenroot, R., Kimonis, E., Hawes, D., Moul, C., Frick, P. J., Anderson, V., & Dadds, M. R. (2017) | Mothers, Fathers, and Parental Systems: A Conceptual Model of Parental Engagement in Programmes for Child Mental Health—Connect, Attend, Participate, Enact (CAPE). | Variabel: Parental Engagement, CAPE model (Connect, Attend, Participate, Enact)<br>Parenting skill, self efficacy, attribution and implementation context. | CAPE model can be used for clinical practice |
| 7 | Rasheed, M. A., Mughis, W., Elahi, K. N., & Hasan, B. S. (2021). | A Family-Centered Intervention to Monitor Children's Development in a Pediatric Outpatient Setting: Design and Feasibility Testing. | Design: quality improvement project<br>Sample: 75,000 patients in pediatric outpatient<br>Variable: family support intervention: the care of child development module, developmental monitoring<br>Instruments: SWYC<br>Analysis: cross sectional descriptive statistics | As many as 54% were detected at risk of cognitive, physical and language disorders. 76% experienced emotional and behavioral disorders. Interventions for family support interventions are easy and acceptable. |
| 8 | Shoghi, M., Shahbazi, B., & Seyedfatemi, N. (2019) | The Effect of the Family-Centered Empowerment Model (FCEM) on the Care Burden of the Parents of Children Diagnosed with Cancer. | Design: quasi-experimental study<br>Sample: 78 parents who have children with cancer<br>Variable: FCEM and care burden<br>Instrument: educational booklet, and parental burden of care.<br>Analysis: Fisher's exact test, chi-square test, independent t-test | The FCEM was implemented in the intervention group in four stages, namely perceived a threat, self-efficacy, educational participation, and evaluation. The results of this study indicate that FCEM reduces the care burden. |
| 9 | Vuorenmaa, M., Perälä, M. L., Halme, N., Kaunonen, M., & Åstedt-Kurki, P. (2016). | Associations between family characteristics and parental empowerment in the family, | Design: cross sectional<br>Sample: 955 parents with children aged 0-9 years<br>Variable: Generic Family Empowerment Scale | Factors that influence the empowerment of mothers and fathers are parental concerns, stress experiences. |
|  |  | family service situations and the family service system. | Analysis: one-way analysis of variance and t-test. |  |
| 10 | Vuorenmaa, Maaret, Halme, N., Perälä, M. L., Kaunonen, M., & Åstedt-Kurki, P. (2016). | Perceived influence, decision-making and access to information in family services as factors of parental empowerment: a cross-sectional study of parents with young children. | Design: cross-sectional<br>Sample: Mothers and fathers of children aged 0-9 years<br>Variables: perceived influence, decision making, access to information in family service. | Design: cross-sectional<br>Sample: Mothers and fathers of children aged 0-9 years<br>Variables: perceived influence, decision making, access to information in family service. |
| 11 | (Bontje et al., 2021) | Parental engagement in preventive youth health care: Effect evaluation | Design: on-randomized controlled trial<br>Sample: Parents with children aged 0-12 years<br>Variable: GIZ (Joint Assessment of Care Needs), concerns discussed, parent-professional agreement on strengths, concerns and follow-up actions and parents' satisfaction<br>Instrument: Questionnaire | The application of GIZ for child health preventive efforts has a positive influence [there are discussions about parenting and environmental conditions as well as parental satisfaction] |
| 12 | Laura Whitehill, Joan Smith, Graham Colditz, Tiffany Le, Polly Kellner, Roberta Pineda<br>(Whitehill et al., 2021) | Socio-demographic factors related to parent engagement in the NICU and the impact of the SENSE program | Design: cross sectional<br>Sample: 70 parents with infant dyads<br>Variables: 1) socio-demographic factors related to parent presence and engagement in the NICU and 2) if the SENSE program (Supporting and Enhancing NICU Sensory Experiences)<br>Instrument: Questionnaire<br>Analysis: 2-Factor ANOVA with a factorial model | Married status, having private insurance, having children are slightly related to good parental involvement<br>The SENSE Program can increase parental engagement in NICU care. |

## Results

Search results and article analysis from the Scopus, Pubmed and Sciencedirect databases, using the PRISMA Flowchart as shown in Figure 1. found 12 articles.

**Figure 1.**
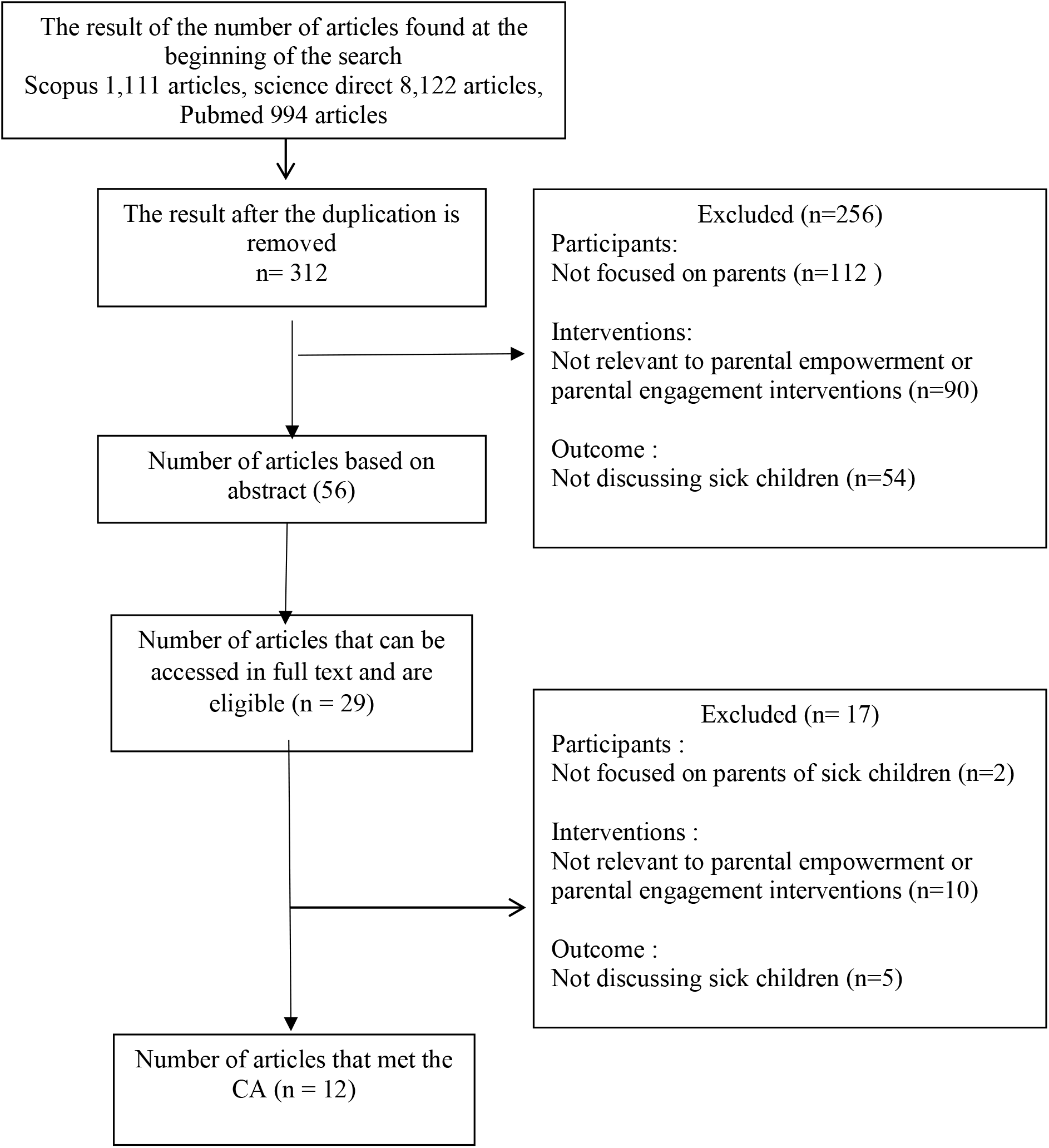
Literature Review flow diagram based on PRISMA

## Discussion

### Ontology of Family Empowerment in Caring Under-Five Children Illness

In essence, children’s health cannot be separated from factors from parents or family, because children are in the system within the family, so the family is a very important element in the growth and development of children, both in healthy and sick children (Yuni Sufyanti Arief et al., 2018). Children have unique characteristics that are different from adults, when children are sick they need specific treatment according to the disease they are experiencing, and the first level of care when a sick toddler is in the family. Families caring for sick toddlers often experience stress caused by worries about the illness experienced by the child and how to treat the child when the child is sick (Mohammadzadeh et al., 2022). Parents caring for sick children are in a state of confusion about the roles they have to do, they tend not to be confident in managing treatment when their children are sick, this results in parents not being able to make the right decisions when the child is sick, even though the role of parents is very important in this condition (Maaret Vuorenmaa et al., 2016).

The discipline of pediatric nursing in improving health and disease prevention in children is oriented toward family-centered nursing services (Family Center Care) which are oriented to capacity building and involving families in every child and family care. The philosophy of pediatric nursing states that the health and nursing services needed by children must involve the family in every process through empowerment (Hockenberry & Wilson, 2013), to prevent trauma, recurrence, and disease severity.

### Epistemological of Family Empowerment in Caring Under-Five Children Illness

Toddlers who are sick are at risk of having problems meeting the needs for growth and development. The family as a unit that is close to the child is expected to be the first level in the care of sick toddlers. This phenomenon is the basic foundation of the importance of developing child nursing science through family empowerment for the care of sick toddlers.

Various studies and theories of family empowerment have been developed as a basis for scientific proof of the philosophy of family-centered nursing. Family-centered nursing is the main intervention for child support. Information obtained by families and children is important in clinical decision-making (Friedman et al., 2003). One of the theories regarding the application of family nursing care with a family-centered nursing approach is the Friedman Model, in which Family Center Nursing is based on the perspective that the family is the basic unit for individual care, the family is the basic unit of a community and society, each of which has its characteristics. different in culture, race, ethnicity, and socioeconomic (Pediatric, 2012).

Treatment with the principles of Family Center Care (FCC) provides family support by increasing self-confidence and the ability to care for children. Two basic concepts in family-centered care are *empowerment* and *enabling* (Hockenberry & Wilson, 2013). FCC provides information from basic to sustainable development, with the aim that families and children can be more effective in care and decision making, providing support facilities both formal and informal. Family empowerment aims to increase knowledge to improve family skills in caring for sick children (Shoghi et al., 2019). The implementation of parental empowerment needs to consider aspects of the personal characteristics of parents and families because the empowerment of parents in caring for children is also related to the personal characteristics of parents or families of sick children. and make decisions, in addition to maternal health, family finances, mothers aged 30 years or older and father’s high education are also related to parental empowerment (M. Vuorenmaa et al., 2016), by looking at the underlying or related parental characteristics, which are expected to realize the appropriate and appropriate application of parental empowerment.

Several studies have stated that family-centered nursing interventions by empowering families are effective in increasing the closeness of children and parents and the ability of parents to care for their children (Rasheed et al., 2021; Arief et al., 2020; Piotrowska et al., 2017), By involving families in child care, it reduces parental concerns about the child care process while the child is sick (M. Vuorenmaa et al., 2016). Another study states that family empowerment by health workers can reduce parental stress and also improve the quality of life of sick children (Mohammadzadeh et al., 2022). The closest family and have the right to make decisions on children including children’s health are parents, both fathers, and mothers, with the application of parental empowerment in health services, they can train parents on how they should play a role in daily child care or when the child is sick, improve their knowledge and skills, can increase their confidence in caring for children so that parents can work together with health workers in making the right decisions in the care of their children (Maaret Vuorenmaa et al., 2016).

WHO recommends one of the treatments for sick toddlers in developing countries through the Integrated Management of Childhood Illness (IMCI), which is an integrated approach to the management of sick toddlers with a focus on the health of children aged 0-5. years (toddlers) as a whole, in this IMCI-based management one of the components is improving family and community practices in-home care and efforts to seek help for sick toddlers (impact increasing community empowerment in health services) (García Sierra & Ocampo Cañas, 2020) (Kementrian kesehatan Republik Indonesia, 2022).

### Axiology of Family Empowerment in Caring Under-Five Children Illness

Children as individuals need to grow and develop optimally. Sick conditions will reduce the quality of life of children and their families, children are at risk of losing the opportunity to get stimulation, carry out normal activities and get nutritional intake that supports their growth and development, so it is important for toddlers who are sick to get the proper care. Nurses play a role in providing nursing care, to achieve the fulfillment of the needs of an individual or family.

The role of nurses is very important to provide appropriate interventions for sick toddlers. A nurse is someone who is highly educated in nursing and carries out nursing activities based on nursing science by providing care to individuals, families, groups, or communities. Family empowerment interventions in sick children based on scientific evidence can be used as the basis for interventions for nurses in the care of sick toddlers (García Sierra & Ocampo Cañas, 2020).

Family Center Care (FCC) is a holistic approach, where the provision of nursing care is not only for individuals (children) but also forms a relationship between nurses and families. (Kuo et al., 2012). The implementation of the FCC focuses on collaboration between nurses and children’s families starting from planning, carrying out, and evaluating actions (Krisnana et al., 2019). The application of FCC aims to involve families in child care, so nurses need to enable families to take care of children when they are sick or healthy (Smeltzer & Bare, 2009). Management of Sick Toddlers through Integrated Management of Childhood Illness (IMCI) in addition to improving the skills of health workers, it also improves family and community practices in-home care and efforts to seek help for sick toddlers (Kementrian kesehatan Republik Indonesia, 2022).

Parental empowerment is a process for parents to better understand the situation, including the condition of a child who is sick (Zimmerman et al., 1992). Family empowerment serves to enable families or improve family competence in interacting with children, through the stages of empowerment and providing assistance to families effectively and efficiently so that families can play a role and make appropriate decisions (Rachmawati et al., 2016).

Family empowerment that can be done by nurses in caring for sick children based on existing theories and research includes providing continuous information, facilitating families with various activities, cooperating in child care, identifying the abilities of children and families, helping to recognize self-ability, increasing the capacity and confidence of children and families (Pediatric, 2012).

Empowerment programs that can be applied based on scientific evidence are through several component approaches such as perceived threats, skills, and self-efficiency, increasing self-confidence through educational participation and evaluation (Shoghi et al., 2019). Another application of family empowerment in caring for sick children with burns is empowerment based on a family-centered care model consisting of four stages, namely threat perception, self-efficacy, self-esteem, and evaluation (Mohammadzadeh et al., 2022). The first stage is the perception of threat, which is to examine the parent’s perception of the child’s health, by explaining the disease that is being experienced by the child starting from the definition, the impact of hospital and home care, and treatment on the treatment plan. The second stage is increasing the self-efficacy of parents, at this stage how a health worker teaches parents how to manage stress, help parents and empower parents in child care. The third stage is self-esteem and self-control, namely by explaining the treatment carried out at home and the healing process and the fourth stage is evaluation, here is an evaluation of what parents have done, evaluation

Research on parental empowerment programs says that parents are agents of change, which in its application says that parental empowerment programs provide support to parents emotionally, help solve problems, help parents develop special competencies, such as effective communication, and help create community with other people. other parents so that they can support each other in the care of their sick children (Olin et al., 2010).

Empowerment of parents or families can be increased by strengthening self-confidence, and providing concrete emotional support so that nurses or health workers can provide better services (M. Vuorenmaa et al., 2016). There are three dimensions in empowering parents in caring for children, namely the family dimension, the dimension of the service situation, and the dimension of the service system. they use, while the service system dimension is the improvement of basic health care services for children. Conceptually, the principle of family empowerment is the same in various places, however, the application can be different, depending on the health service system used and conditions in the local area (M. Vuorenmaa et al., 2016).

According to M. Vuorenmaa et al., (2016) success in parental empowerment in the family aspect is seen from the aspect of fathers and mothers, fathers and mothers who are confident in their skills in parenting, do not experience difficulties when caring for children, and feel that there is easy access when they need help. Indicators of increasing maternal empowerment in situations include when the mother feels confident in her skills and responsibilities as a parent in caring for her child, does not feel worried about the child’s physical health, and does not find it difficult to take care of her family or carry out work, feel easy to access services. 30 years and a place for daily child care, while in the aspect of the indicator paragraph in increasing father empowerment, namely when fathers believe in child care skills, do not find it difficult to work and take care of their daily families, and facilitate child care services. In the dimension of the family service system, indicators of increasing maternal empowerment in child care are confidence in their child-rearing skills and roles as parents, not finding it difficult to take care of their family and carry out daily work, and being 30 years of age or older and where children are cared for daily, while from the father’s factor the indicators of success in increasing parental empowerment are the father’s confidence in his child-rearing skills, ease of access to assistance and the high level of father education.

## Conclusion

Sick toddlers need proper care and treatment to prevent the severity of the disease that can have an impact on disrupting the growth and development process of Toddlers, the family is the closest unit to under-five children who can determine the Toddler’s health. A philosophical review of family empowerment in the care of sick children can be the basis for pediatric nurses in providing nursing care to sick toddlers. Nurses can implement women’s empowerment programs by providing continuous information, facilitating families with various activities, cooperating in child care, identifying the abilities of children and families, helping to recognize self-ability, and increasing the capacity and confidence of children and families.

## Data Availability

All data produced in the present study are available upon reasonable request to the authors

## Reference

Arief, Y S Nursalam, & Rachmawati, P. D. (2020). Family filial value on caring children with leukemia. Enfermeria Clinica, 30, 157–160. https://doi.org/10.1016/j.enfcli.2019.11.044

Arief, Yuni Sufyanti, Nursalam, N., Ugrasena, I. D. G., Devy, S. R., & Savage, E. (2018). The Development of Model Family-Centered Empowerment on Caring for Children with Leukemia. Jurnal Ners, 13(1), 98. https://doi.org/10.20473/jn.v13i1.7774

Bontje, M. C. A., de Ronde, R. W., Dubbeldeman, E. M., Kamphuis, M., Reis, R., & Crone, M. R. (2021). Parental engagement in preventive youth health care: Effect evaluation. Children and Youth Services Review, 120, 105724. https://doi.org/10.1016/j.childyouth.2020.105724

Damen, H., Scholte, R. H. J., Vermulst, A. A., van Steensel, P., & Veerman, J. W. (2021). Parental empowerment as a buffer between parental stress and child behavioral problems after family treatment. Children and Youth Services Review, 124, 105982. https://doi.org/10.1016/j.childyouth.2021.105982

Friedman, M. M., Bowden, V. R., & Jones, E. (2003). Family nursing: Research, theory & practice (Vol. 16). Prentice Hall Upper Saddle River, NJ.

García Sierra, A. M., & Ocampo Cañas, J. A. (2020). Integrated management of childhood illnesses implementation-related factors at 18 colombian cities. BMC Public Health, 20(1), 1–10. 10.1186/S12889-020-09216-0/TABLES/6

Hockenberry, M. J., & Wilson, D. (2013). Wong’s Esential of Pediatric Nursing (9th ed.). Elsevier Mosby.

Kementrian kesehatan Republik Indonesia. (2022). Buku Bagan Manajemen Terpadu Balita Sakit. Kementrian Kesehatan Republik Indonesia.

Krisnana, I., Sulistyarini, H., Rachmawati, P. D., Arief, Y. S., & Kurnia, I. D. (2019). Reducing acute stress disorders in mothers of leukemic children by means of the family Centered Empowerment Module (FACE). Central European Journal of Nursing and Midwifery, 10(2), 1035–1040. 10.15452/CEJNM.2019.10.0011

Kuo, D. Z., Houtrow, A. J., Arango, P., Kuhlthau, K. A., Simmons, J. M., & Neff, J. M. (2012). Family-Centered Care: Current Applications and Future Directions in Pediatric Health Care. Maternal and Child Health Journal, 16(2), 297–305. 10.1007/s10995-011-0751-7

May, M., Brousseau, D. C., Nelson, D. A., Flynn, K. E., Wolf, M. S., Lepley, B., & Morrison, A. K. (2018). Why Parents Seek Care for Acute Illness in the Clinic or the ED: The Role of Health Literacy. Academic Pediatrics, 18(3), 289–296. 10.1016/J.ACAP.2017.06.010

Mohammadzadeh, E., Varzeshnejad, M., Masoumpour, A., & Ahmadimehr, F. (2022). The impact of the family-centered empowerment model on the children’s quality of life with chemical burns and their parent’s perceived stress. Burns. https://doi.org/10.1016/j.burns.2022.06.002

Olin, S. S., Hoagwood, K. E., Rodriguez, J., Ramos, B., Burton, G., Penn, M., Crowe, M., Radigan, M., & Jensen, P. S. (2010). The Application of Behavior Change Theory to Family-Based Services: Improving Parent Empowerment in Children’s Mental Health. Journal of Child and Family Studies, 19(4), 462–470. 10.1007/s10826-009-9317-3

Pediatric, A. A. O. (2012). Patient- and Family-Centered Care and the Pediatrician’s Role. Pediatrics, 129(2), 394–404. www.pediatrics.org

Piotrowska, P. J., Tully, L. A., Lenroot, R., Kimonis, E., Hawes, D., Moul, C., Frick, P. J., Anderson, V., & Dadds, M. R. (2017). Mothers, Fathers, and Parental Systems: A Conceptual Model of Parental Engagement in Programmes for Child Mental Health— Connect, Attend, Participate, Enact (CAPE). Clinical Child and Family Psychology Review, 20(2), 146–161. 10.1007/s10567-016-0219-9

Rachmawati, P. D., Kurnia, I. D., Asih, M. N., Kurniawati, T. W., Krisnana, I., Arief, Y. S., Mani, S., Dewi, Y. S., & Arifin, H. (2022). Determinants of under-five mortality in Indonesia: A nationwide study. Journal of Pediatric Nursing, 65, e43–e48. 10.1016/j.pedn.2022.02.005

Rachmawati, P. D., Ranuh, R., & Arief, Y. (2016). Model Pengembangan Perilaku Ibu dalam Pemenuhan Kebutuhan Asah Asih dan Asuh pada Anak dengan Leukemia. Jurnal NERS, 11(1), 63. 10.20473/jn.V11I12016.63-72

Rasheed, M. A., Mughis, W., Elahi, K. N., & Hasan, B. S. (2021). A Family-Centered Intervention to Monitor Children’s Development in a Pediatric Outpatient Setting: Design and Feasibility Testing. Frontiers in Health Services, 0, 6. 10.3389/FRHS.2021.739655

Shoghi, M., Shahbazi, B., & Seyedfatemi, N. (2019). The Effect of the Family-Centered Empowerment Model (FCEM) on the Care Burden of the Parents of Children Diagnosed with Cancer. Asian Pacific Journal of Cancer Prevention : APJCP, 20(6), 1757–1764. 10.31557/APJCP.2019.20.6.1757

Smeltzer, S. C., & Bare, B. G. (2009). Buku Ajar Keperawatan Medikal Bedah Brunner & Suddarth. EGC.

USAID. (2020). The DHS Program Demographic and Health Surveys. https://dhsprogram.com/

Vuorenmaa, M., Perälä, M. L., Halme, N., Kaunonen, M., & Åstedt-Kurki, P. (2016). Associations between family characteristics and parental empowerment in the family, family service situations and the family service system. Child: Care, Health and Development, 42(1), 25–35. 10.1111/CCH.12267

Vuorenmaa, Maaret, Halme, N., Perälä, M. L., Kaunonen, M., & Åstedt-Kurki, P. (2016). Perceived influence, decision-making and access to information in family services as factors of parental empowerment: a cross-sectional study of parents with young children. Scandinavian Journal of Caring Sciences, 30(2), 290–302. 10.1111/SCS.12243

Whitehill, L., Smith, J., Colditz, G., Le, T., Kellner, P., & Pineda, R. (2021). Socio-demographic factors related to parent engagement in the NICU and the impact of the SENSE program. Early Human Development, 163, 105486. https://doi.org/10.1016/j.earlhumdev.2021.105486

Zimmerman, M. A., Israel, B. A., Schulz, A., & Checkoway, B. (1992). Further explorations in empowerment theory: An empirical analysis of psychological empowerment. American Journal of Community Psychology 1992 20:6, 20(6), 707–727. 10.1007/BF01312604

